## Supplementary material for "Menopause education of healthcare professionals: A scoping review protocol": S1

**PRISMA-P (Preferred Reporting Items for Systematic Review and Meta-Analysis Protocols) 2015 Checklist**

*Recommended items to address in a systematic review protocol*

| **Section and Topic** | **Item No** | **Checklist Item** | **Page number** |
| --- | --- | --- | --- |
| **ADMINISTRATIVE INFORMATION** |  |  |  |
| **Title** | 1a | Identify the report as a protocol of a systematic review | 1 |
|  | 1b | If an update, identify as such | N/A |
| **Registration** | 2 | Provide registry name (e.g., PROSPERO) and registration number | TBC |
| **Authors** | 3a | Names, affiliations, emails, and mailing address of corresponding author | 1 |
|  | 3b | Describe author contributions and identify review guarantor | N/A |
| **Amendments** | 4 | Document protocol amendments or state plan for recording changes | 8 |
| **Support** | 5a | Sources of financial or other support | 1 |
|  | 5b | Name of funder/sponsor | 1 |
|  | 5c | Role of funder/sponsor in protocol development | N/A |
| **INTRODUCTION** |  |  |  |
| **Rationale** | 6 | Describe review rationale in context of existing knowledge | 2,3,4 |
| **Objectives** | 7 | Explicit statement of review questions (PICO) | 4,5 |
| **METHODS** |  |  |  |
| **Eligibility Criteria** | 8 | Specify study/report characteristics (PICO, design, timeframe, language, etc.) | 4,5,6,7,8 |
| **Information Sources** | 9 | List databases, grey literature, and planned coverage dates | 2,6 |
| **Search Strategy** | 10 | Draft search strategy for at least one database (with reproducible limits) | 2, S1 Appendix: Search strategy |
| **Study Records** | 11a | Data management methods (e.g., Covidence, Ref 24) | 2,7 |
|  | 11b | Study selection process (e.g., independent dual screening) | 7 |
|  | 11c | Data extraction method (e.g., piloted forms, dual extraction) | 7 |
| **Data Items** | 12 | List and define all variables (PICO, assumptions) | 5,6 |
| **Outcomes & Prioritization** | 13 | Define and prioritize outcomes with rationale | 9 |
| **Risk of Bias** | 14 | Methods for assessing bias (study/outcome level) and use in synthesis | N/A |
| **Data Synthesis** | 15a | Criteria for quantitative synthesis | N/A |
|  | 15b | Planned summary measures, data handling, and combination methods (e.g., I²) | 5 |
|  | 15c | Additional analyses (subgroups, sensitivity, meta-regression) | N/A |
|  | 15d | Summary plan if quantitative synthesis is inappropriate | 8 |
| **Meta-Bias(es)** | 16 | Assessment of publication/selective reporting bias | N/A |
| **Confidence in Evidence** | 17 | Method to evaluate evidence strength (e.g., GRADE) | N/A |

*** It is strongly recommended that this checklist be read in conjunction with the PRISMA-P Explanation and Elaboration (cite when available) for important clarification on the items. Amendments to a review protocol should be tracked and dated. The copyright for PRISMA-P (including checklist) is held by the PRISMA-P Group and is distributed under a Creative Commons Attribution Licence 4.0.**

*From: Shamseer L, Moher D, Clarke M, Ghersi D, Liberati A, Petticrew M, Shekelle P, Stewart L, PRISMA-P Group. Preferred reporting items for systematic review and meta-analysis protocols (PRISMA-P) 2015: elaboration and explanation. BMJ. 2015 Jan 2;349(jan02 1):g7647.*
