## Supplementary material for "Menopause education of healthcare professionals: A scoping review protocol": S2

S2 Appendix: Data extraction table 1

| **Authors/**  **Year** | **Country** | **Setting** | **Study Design** | **Population Type** | **Population Size** | **Study aims** | **Methodology** |
| --- | --- | --- | --- | --- | --- | --- | --- |
