## Supplementary material for "Menopause education of healthcare professionals: A scoping review protocol": S3

S3 Appendix: Data extraction table 2

| **Authors/**  **Year** | **Intervention** | **Facilitators** | **Participants/**  **Profession** | **Course delivery method** | **Course length** | **Course outline** |
| --- | --- | --- | --- | --- | --- | --- |
