## Supplementary material for "Menopause education of healthcare professionals: A scoping review protocol": S4

S1 Appendix: Medline search strategy

| Concept 1  **Menopause**  (n= 79,708) | Concept 2  **Education**  (n= 5,639,663 ) | Concept 3  **Healthcare Professionals**  (n= 2,146,884 |
| --- | --- | --- |
| TI ( menopause OR menopausal OR Perimenopause or perimenopausal or climacteric )  OR  AB ( menopause OR menopausal or perimenopause OR perimenopausal OR climacteric )  OR  (MM "Menopause")  OR  (MM "Climacteric+")  OR  (MM "Perimenopause")  OR  (MM "Postmenopause") OR  (MM "Menopause, Premature") | TI ( education OR learning or teaching OR “education* system*” OR curriculum OR curricula OR pedagogy OR pedagogical) OR undergrad* OR postgrad*)  OR  (MM "Education, Continuing+")  OR  (MM "Education, Distance")  OR  (MM "Education+")  OR  (MM "Health Education+")  OR  (MM "Teaching Materials+")  OR  (MM "Hospitals, Teaching+") | TI ( Physicians, Primary Care") OR "nurs* OR physician* OR doctor* OR midwi* OR pharmac* OR physiotherap* OR psycholog" )  OR  AB ( Physicians, Primary Care") OR "nurs* OR physician* OR doctor* OR midwi* OR pharmac* OR physiotherap* OR psycholog" )  OR  (MM "Students, Nursing") OR "student nurses OR nursing students or student nurse OR nursing student OR undergraduate nurse"  OR  (MM "Students, Medical") OR (MM "Education, Medical+") OR (MM "Education, Medical, Graduate+") OR (MM "Education, Medical, Continuing") OR (MH "Medicine") OR medicine students OR students in medicine OR medical education OR medical studies OR residents" OR (MM "Education, Medical, Undergraduate")  OR  MM "Primary Health Care+") OR (MM "General Practice+") OR "general practice OR GP OR doctor or primary care" OR (MM "Primary Nursing") OR (MM "Primary Care Nursing") OR (MM "General Practitioners") OR (MM "Physicians, Primary Care") OR (MM "Physicians+") OR (MM "Physicians, Women") OR (MM "Physicians, Family") OR (MM "Practice Patterns, Physicians'")  OR(MH "Nurse Midwives") OR (MM "Nurses+") OR (MM "Nurses, Community Health") OR (MM "Nurses, Public Health") OR "nurse or midwife"  OR  (MM "Practice, Psychological") |
